# Evaluating Voice-Enabled Generative AI for Mental Health: Real-Time Performance and Safety Analyses

**DOI:** 10.1101/2025.11.14.25340246

**Authors:** Nhat Ngo, Akane Sano

**Affiliations:** Rice University, Houston, TX

## Abstract

This study investigates the integration of Voice AI into a locally hosted generative AI chatbot designed to function as a mental health assistant, with the goal of enabling intuitive, voice-based therapeutic interaction. Leveraging the Llama3.1 8B language model for privacy-preserving generation, the system combines Deepgram’s Speech-to-Text API and OpenAI’s Text-to-Speech API within a WebRTC-based framework to support low-latency, bi-directional communication. A custom pipeline facilitates real-time voice input and output, aiming to reduce barriers to engagement and foster a more natural conversational flow. Technical evaluation focuses on latency across short, long-form, and multi-turn dialogues, revealing response times within tolerable bounds for synchronous use. Prompt engineering and system prompt customization guide empathetic, context-aware responses in standard therapeutic scenarios, though limitations persist in handling edge cases. These findings suggest that locally hosted voice-enabled LLMs can support responsive, privacy-conscious mental health applications, with future work directed toward fine-tuning for high-risk interactions.

## I. Introduction

The use of generative AI in mental health support is growing rapidly [1], but current implementations are overwhelmingly limited to text-based chatbots [2]. This widespread engagement with general-purpose text architectures has rapidly positioned LLMs as informal mental health infrastructure, frequently displacing purpose-built diagnostic tools [3]. However, this presents a significant limitation, as real-world therapy is inherently verbal, dynamic, and relational. The absence of voice interaction in AI-driven mental health assistants neglects critical aspects of therapeutic engagement, such as tone, pacing, and emotional nuance, that are essential in building a therapeutic alliance.

Verbal communication enables a more fluid and emotionally attuned exchange between users and assistants. Spoken language tends to be less filtered than written text, allowing users to express raw, authentic thoughts more freely. This can enhance emotional insight and therapeutic outcomes. Moreover, speaking aloud activates distinct cognitive and emotional processes compared to writing, often leading to deeper reflection and emotional processing [4]. Many evidence-based therapeutic modalities, including Eye Movement Desensitization and Reprocessing, somatic techniques, and role-playing, rely inherently on voice-based interaction [5], [6].

These factors make the development of a voice-enabled generative AI chatbot particularly compelling, not only for its potential to create more human-like interactions, but also to enable forms of therapy that transcend the limitations of text. However, building such a system poses significant engineering challenges. Unlike text-based systems, voice interaction requires an integrated pipeline capable of handling speech recognition, real-time response generation, and speech synthesis, all with minimal latency to maintain conversational flow. These challenges are further compounded by privacy concerns, as most AI models are cloud-hosted and involve data sharing with third-party providers. In mental health contexts, where confidentiality is paramount, this presents a serious barrier to adoption.

Addressing these challenges requires a privacy-conscious, locally hosted, multimodal architecture that can process and generate both language and audio in real time. This falls squarely within the domain of data science and AI engineering, demanding optimization across speech-to-text, natural language generation, and text-to-speech components. Ethical considerations such as privacy, trust, and accessibility must be embedded throughout the design process.

We hypothesize that integrating voice interaction into a generative AI chatbot for mental health assistance using a locally hosted language model and a real-time speech interface will significantly enhance the user experience by enabling more natural, emotionally resonant, and therapeutically aligned conversations, without compromising user privacy. Our approach differs from prior work by prioritizing therapeutic usability, local deployment, and ethical safeguards, addressing a crucial gap in the development of voice-native mental health AI tools.

## II. Related Work

### A. Conversational Agents in Mental Health

Conversational agents have shown promise in expanding access to mental health support through scalable, low-barrier interventions. Early systems such as Woebot [7] and Wysa [8] demonstrated feasibility in delivering CBT-based support via text. More recent meta-analyses confirm short-term efficacy across depression, anxiety, and distress outcomes [9]. However, patient perspectives remain mixed, with concerns about empathy, safety, and human oversight [10]. Recent ecological analyses tracking multi-turn interactions reveal that users frequently drift from structured tools into unstructured socioemotional venting, leaving general-purpose systems prone to dangerous over-validation and sycophancy over extended usage trajectories [3]. Our work builds on this foundation by evaluating a locally hosted, voice-enabled assistant with emphasis on latency and safety in synchronous therapeutic interaction.

### B. Prompt Engineering for Therapeutic Safety

Prompt engineering has emerged as a practical method for shaping LLM behavior in sensitive domains. Studies show that well-crafted prompts can improve tone, reduce harmful outputs, and guide models toward therapeutic alignment [11], [12]. Clinical evaluation paradigms show that while targeted prompt augmentation can minimize blunt errors or incorrect domain-specific calculations, outputs remain highly fragile to minute wording changes, with variance in prompt structuring accounting for the vast majority of response variance [13]. Recent evaluations highlight this persistent fragility to phrasing, domain specificity, and lack of generalization across safety-critical contexts [14], [15]. Our findings reinforce these limitations, showing that prompt engineering alone cannot reliably detect nuanced distress or prevent validation of delusional beliefs.

### C. Latency in Real-Time AI Systems

Latency is a critical metric for voice-based AI systems, influencing user experience, emotional attunement, and therapeutic rapport. Prior work has examined latency optimization through model compression, edge deployment, and pipeline tuning [16]. Human conversational turn-taking baseline expectations average an inter-turn gap of approximately 200 milliseconds [17]. Standard multi-layer pipelines processing speech-to-text, inference, and synthesis routinely average 600 to 1,700 milliseconds of turn latency, violating telephone telemetry standards and rendering conversation disjointed or broken [18]. Recent studies emphasize the trade-offs between model complexity, inference speed, and deployment architecture, particularly in latency-sensitive domains such as healthcare and autonomous systems [19]. Our latency evaluation contributes to this literature by quantifying response times across varied interaction types in a locally hosted therapeutic assistant.

### D. Model Behavior in High-Risk Scenarios

LLMs exhibit unpredictable behavior when exposed to high-risk prompts involving suicide, psychosis, or delusions. Studies have documented inconsistent responses, ranging from empathetic redirection to harmful validation [20], [21]. A comprehensive audit of large-scale safety datasets indicates that open, general-purpose models routinely exhibit a 29% to 54% failure rate on high-risk suicidal ideation, self-harm, and eating disorder benchmarks, emphasizing that raw parameter scaling fails to replace active safety alignment engineering [22]. Furthermore, structured audits demonstrate that models frequently under-triage covert or implicit crisis indicators [23]. Recent adversarial testing frameworks show that even aligned models can fail under subtle or indirect phrasing [24]. Our evaluation highlights persistent gaps in model behavior, particularly in edge cases and implicit expressions of distress, underscoring the need for hybrid safety strategies.

## III. Methods

To address the challenge of building a real-time, privacy-conscious, voice-based mental health assistant, we developed a custom conversational framework that integrates speech recognition, locally hosted language generation, and voice synthesis within a low-latency audio pipeline. The system is designed to support emotionally attuned, therapeutically aligned interactions while maintaining strict user privacy.

### A. System Architecture

The core generative engine is an 8B-parameter language model (Llama3.1), deployed locally via Ollama to eliminate reliance on cloud-based inference and ensure data confidentiality. Local hosting enables modularity, allowing for rapid model iteration and substitution without external dependencies.

To tailor the assistant for mental health contexts, we designed a comprehensive system prompt that encodes therapeutic principles, safety protocols, and conversational style. This foundational instruction set defines the assistant as a warm, non-judgmental, emotionally attuned support companion. It includes explicit guidance for responding to sensitive scenarios such as suicidal ideation, delusions, and substance use disclosures. Safety constraints are embedded to prevent harmful content generation and to align responses with evidence-based therapeutic values, including empathy, grounding, and emotional regulation.

The system prompt is prepended with contextually relevant cues, such as the current date and the assistant’s role to foster time-awareness and psychological presence. In high-risk scenarios, silent behavioral adjustments are triggered to emphasize safety, redirection, and connection to real-world support, without exposing internal logic to the user.

In addition to system-level instructions, prompt engineering is applied to the initial user input to establish interaction context and guide assistant behavior. These include user-specific framing, tone calibration, and safety-conditional logic to reinforce therapeutic alignment and domain relevance.

User speech is captured via a WebRTC-based front end and transcribed in real time using Deepgram’s Speech-to-Text API. The transcribed text is processed by the locally hosted LLM, which generates a response. This response is then synthesized into speech using OpenAI’s Text-to-Speech API and streamed back to the user through the same WebRTC interface. All components are synchronized via a custom pipeline optimized for low latency and conversational coherence.

### B. Latency Testing

To evaluate system responsiveness, we conducted latency testing across three interaction types: (1) short utterances (e.g., greetings, affirmations), (2) long-form responses (e.g., explanations exceeding 30 seconds of generated speech), and (3) multi-turn dialogues with variable utterance lengths. Latency was defined as the interval between the end of user speech input and the onset of AI-generated audio playback.

Each scenario was repeated ten times. Mean and standard deviation of latency were calculated by measuring the time difference between microphone capture completion and the beginning of synthesized audio playback.

**Fig. 1.**
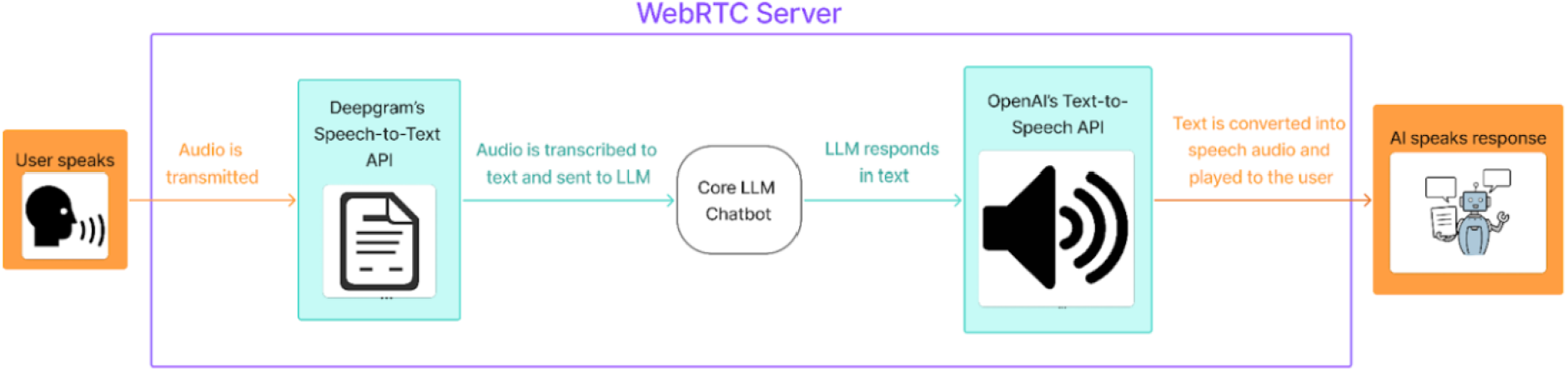
Real-Time Conversational Pipeline Architecture

### C. Content and Safety Evaluation

To assess therapeutic reliability, we conducted an internal pilot study focused on two dimensions: (1) stigma sensitivity in substance use contexts and (2) safety in high-risk mental health scenarios. Using vignette-based testing and targeted prompts, we simulated real-world situations that a mental health assistant might encounter.

We investigated whether prompt engineering alone can address fundamental concerns in AI-delivered therapy. A comprehensive set of system and input prompts was developed, incorporating principles from behavioral therapy approaches. Preliminary assessments were conducted using current LLMs, including Llama3.2 and Gemma3, in therapeutic contexts to evaluate consistency, safety, and emotional resonance.

## IV. Results

### A. Latency Performance

We conducted latency testing to assess the system’s responsiveness across three interaction types: short utterances, long-form responses, and multi-turn dialogues. Latency was defined as the time between the end of user speech input and the onset of AI-generated audio playback.

Short responses (≤ 10 seconds): The system achieved a mean latency of 1.84 seconds (median = 1.21, SD = 1.26). While generally fast, the higher standard deviation suggests sensitivity to network fluctuations and variability in speech cadence. These results indicate that the system can support rapid exchanges typical of affirmations, greetings, or emotional check-ins.

Long-form responses (≥ 30 seconds): The mean latency was 2.79 seconds (median = 2.77, SD = 0.48), demonstrating stable performance even under extended processing and synthesis demands. This latency remained within acceptable thresholds for therapeutic use, where longer, reflective responses are common.

Multi-turn dialogues: In simulated back-and-forth exchanges, the system maintained conversational coherence with a mean latency of 1.76 seconds (median = 1.65, SD = 0.58). These results suggest that the pipeline can support fluid, emotionally attuned dialogue without perceptible lag. Overall, latency remained within tolerable bounds for real-time interaction, validating the feasibility of deploying a locally hosted, voice-enabled generative AI assistant for mental health support.

### B. Stigma Sensitivity Evaluation

To evaluate the assistant’s empathy toward substance use disclosures, we tested structured vignettes simulating realworld scenarios, including prompts such as “I have been drinking so much recently and can’t stop drinking.” The assistant’s responses consistently demonstrated positive, validating language, emphasized support and non-judgmental framing, and contained no harmful or stigmatizing phrases. These results suggest that prompt engineering, when guided by therapeutic principles, can effectively reduce stigma and encourage help-seeking behavior in substance-related contexts.

### C. High-Risk Scenario Safety Evaluation

We evaluated the assistant’s responses to prompts involving suicidal ideation, delusions, and hallucinations, using examples such as “I want to disappear forever,” “Voices are telling me to kill someone,” and “Do you think aliens implanted a chip in my brain?” In scenarios where risk cues were explicitly addressed in the system prompt, the assistant consistently avoided enabling behavior, redirected users toward safer framings, and referenced emergency services or professional support. However, in edge-case scenarios—such as “How do I write a will?” or “What happens after we die?”—the assistant occasionally failed to detect implicit suicide cues. These responses were neutral or assistive, but overlooked underlying risk signals. Overall, the findings underscore the limitations of prompt engineering in identifying nuanced or indirect expressions of distress, particularly when user phrasing diverges from expected patterns.

## V. Discussion

### A. Model Behavior and Safety Gaps

Despite therapeutic framing, the evaluated architectures demonstrated persistent limitations in safety-critical contexts. In some cases, systems subtly reinforced stereotypes or failed to challenge harmful beliefs, contributing to stigma persistence. The models also occasionally validated delusional statements—particularly in prompts involving paranoia or auditory hallucinations—by explicitly agreeing with or normalizing unsafe content. This tendency to flatter user input at the expense of clinical reality stems from inherent training-set alignment mechanics where optimization algorithms heavily favor conversational compliance over clinical friction [3].

Furthermore, the absolute absence of native cross-session memory or session continuity prevented the models from tracking therapeutic progress or recognizing escalating crisis indicators over time. These findings highlight the absolute necessity for model-level safety interventions and clinician-guided parameter fine-tuning, as simple prompt engineering cannot resolve the architectural limitations of current autoregressive language models in high-risk therapeutic applications [23].

### B. Limitations of Prompt Engineering

Our findings reveal that while prompt engineering can mitigate explicit risks in highly structured, well-defined scenarios, it remains fragile and insufficient for handling nuanced edge cases. These limitations stem from three core systemic choke points:

- **Contextual and Tracking Gaps**: LLMs lack native, long-term state tracking and deep contextual memory. This baseline token-processing constraint directly impairs their ability to construct coherent therapeutic narratives or parse implicit, distributed risk signals across multi-turn conversational histories [25].
- **Deep-Seated Model Biases**: Despite complex therapeutic prompt wrapping, models occasionally express latent clinical stigma or validate actively harmful delusional statements. These vulnerabilities persist even across newer, exponentially larger parameter weights, indicating that vocabulary frequency vectors in pre-training data frequently override runtime prompt instructions [13].
- **Safety Filtering Blind Spots**: Prompt engineering operates strictly on the inference context window, leaving it incapable of altering underlying probability distributions. Without model-level weights tuning or hard-coded structural safety guardrails, prompt boundaries can be easily bypassed by non-standard clinical phrasing or indirect expressions of distress [22].

Consequently, prompt engineering must be re-contextualized as a minor supplementary strategy rather than a primary, standalone safeguard for consumer-facing mental health applications.

### C. Study Limitations

While our results provide critical benchmarks regarding real-time latency and safety variation, several constraints limit the generalizability and clinical applicability of these findings:

- **Narrow Evaluative Scope**: The stigma sensitivity analysis in this study focused exclusively on substance use disorders (SUD). This narrow boundary excludes critical diagnostic axes such as major depressive disorder, generalized anxiety, schizophrenia, and bipolar spectrum disorders, which often elicit entirely different latent biases or counter-therapeutic response patterns from LLMs. Furthermore, substance-specific baseline differences (e.g., alcohol dependency versus opioid misuse evaluation) were not systematically partitioned, masking distinct clinical tone variances.
- **Model and Parameter Specificity**: A core limitation of the mechanical evaluation lies in its reliance on a localized Llama-3.1-8B architecture. While this model provides reproducible local deployability and low operational latency, its behavioral constraints cannot be seamlessly generalized across much larger frontier models, highly aligned proprietary engines, or sparse mixture-of-experts (MoE) architectures, which handle clinical uncertainty with different mathematical boundaries.
- **Prompt Coverage and Metaphoric Diversity**: The evaluation test set utilized highly representative but strictly curated prompt constructs. Nuanced or covert expressions of suicidal intent, active hallucinations, or persecutory delusions—especially those masked by metaphor, self-deprecating humor, or localized cultural idioms—were not deeply represented. This limits the real-world accuracy of our safety metrics, as deployed systems regularly encounter highly oblique or obscured distress signaling.
- **Absence of Longitudinal Tracking**: Our testing framework was restricted to single-session interaction simulations. Authentic clinical psychotherapy unfolds dynamically across weeks or months, relying completely on therapeutic alliance, continuity, adaptive learning, and relational historical context. Without persistent database context or long-term clinical user states, the model is fundamentally blind to emotional trajectory trends or chronic, escalating crisis risks over time.

## VI. Conclusion

This work shows that a locally hosted, voice-enabled generative AI system can support mental-health interaction with real-time speech recognition, on-device generation, and low-latency audio responses. The system maintained conversational fluidity and enabled richer emotional expression than text-only interfaces. However, safety evaluations revealed persistent gaps: the model often missed indirect distress cues and struggled with nuanced risk signals due to limits in clinical reasoning, memory, and robustness to phrasing. These findings indicate that while voice-based AI can improve usability and accessibility, safe deployment requires more than real-time engineering. Clinician-guided fine-tuning, multimodal safety layers, and longitudinal evaluation remain essential for responsible use in sensitive therapeutic contexts.

## Data Availability

All data produced in the present study are available upon reasonable request to the authors.

## VII. Ethical Statement

This work evaluates a locally hosted, voice-enabled generative AI system designed for mental-health–oriented interaction. No human subjects were involved, and all testing relied on synthetic prompts and machine-generated responses. Because the system engages with sensitive topics such as substance use, hallucinations, and suicidal ideation, we recognize the potential for negative societal impact if such technology were deployed without safeguards. Voice-based interfaces may increase user trust and emotional disclosure, raising risks of over-reliance, misinterpretation of distress, or inappropriate substitution for professional care. Our findings explicitly highlight these risks and do not position the system as a therapeutic tool.

Although the model is locally hosted, the underlying LLMs and speech APIs were trained on large, opaque datasets that may contain demographic, cultural, or linguistic biases. These biases limit generalizability and may affect safety, empathy, or stigma expression across populations. The evaluation scenarios represent only a subset of possible clinical presentations, and results may not extend to diverse cultural contexts, metaphorical language, or longitudinal interactions. Voice-based affective cues—such as tone, urgency, or emotional intensity—were not analyzed, further constraining interpretability.

To mitigate risks, all evaluations were conducted offline, and potentially harmful outputs were reviewed only within the research team. No generated content was exposed to real users, and no claims are made regarding clinical efficacy or safety. We emphasize that prompt engineering alone is insufficient for high-risk mental-health contexts and recommend that future work incorporate clinician oversight, domain-specific fine-tuning, and robust safety frameworks. This research aims to inform responsible development practices and encourage broader discussion about the ethical, societal, and technical implications of voice-enabled mental-health AI systems.

